# Mitochondrial DNA heteroplasmy distinguishes disease manifestation in *PINK1*- and *PRKN*-linked Parkinson’s disease

**DOI:** 10.1101/2022.05.17.22275087

**Authors:** Joanne Trinh, Andrew A. Hicks, Inke R. König, Sylvie Delcambre, Theresa Lüth, Susen Schaake, Kobi Wasner, Jenny Ghelfi, Max Borsche, Carles Vilariño-Güell, Faycel Hentati, Elisabeth L. Germer, Peter Bauer, Masashi Takanashi, Vladimir Kostić, Anthony E. Lang, Norbert Brüggemann, Peter P. Pramstaller, Irene Pichler, Alex Rajput, Nobutaka Hattori, Matthew J. Farrer, Katja Lohmann, Hansi Weissensteiner, Patrick May, Christine Klein, Anne Grünewald

**Author notes:** Corresponding Author: Anne Grünewald, PhD, Luxembourg Centre for Systems Biomedicine, University of Luxembourg, Campus Belval, 6, avenue du Swing, L-4367 Belvaux, Luxembourg.

## Abstract

Biallelic mutations in *PINK1* and PRKN cause recessively inherited Parkinson’s disease (PD). Though some studies suggest that *PINK1*/*PRKN* monoallelic mutations may not contribute to risk, deep phenotyping assessment showed that *PINK1* or *PRKN* monoallelic pathogenic variants were at a significantly higher rate in PD compared to controls. Given the established role of *PINK1* and Parkin in regulating mitochondrial dynamics, we explored mitochondrial DNA (mtDNA) integrity and inflammation as potential disease modifiers in carriers of mutations in these genes. MtDNA integrity, global gene expression and serum cytokine levels were investigated in a large collection of biallelic (n=84) and monoallelic (n=170) carriers of *PINK1*/*PRKN* mutations, iPD patients (n=67) and controls (n=90). Affected and unaffected *PINK1*/*PRKN* monoallelic mutation carriers can be distinguished by heteroplasmic mtDNA variant load (AUC=0.83, CI:0.74-0.93). Biallelic *PINK1*/*PRKN* mutation carriers harbor more heteroplasmic mtDNA variants in blood (p=0.0006, Z=3.63) compared to monoallelic mutation carriers. This enrichment was confirmed in iPSC-derived and postmortem midbrain neurons from biallelic *PRKN*-PD patients. Lastly, the heteroplasmic mtDNA variant load was found to correlate with IL6 levels in *PINK1*/*PRKN* mutation carriers (r=0.57, p=0.0074). *PINK1*/*PRKN* mutations predispose individuals to mtDNA variant accumulation in a dose- and disease-dependent manner. MtDNA variant load over time is a potential marker of disease manifestation in *PINK1*/*PRKN* mutation carriers.

## Introduction

Parkinson’s disease (PD) is a neurodegenerative disorder affecting over seven million patients world-wide[9], of which ∼10% can be genetically explained [3, 9]. Multiple genes have been found to be implicated in familial and sporadic PD, including pathogenic mutations and strong risk factors. Biallelic *PINK1* or *PRKN* mutations are a well-known cause of early-onset PD [15, 37]. Interestingly, monoallelic *PINK1*/*PRKN* mutations have been considered a PD risk factor [20]. Based on a conservative estimate of a frequency of 2% of monoallelic *PINK1* and *PRKN* mutations among all PD patients, one can expect ∼140,000 carriers worldwide, this warrants a deeper understanding of the possible functional consequence of *PINK1* or *PRKN* monoallelic mutations. Affected monoallelic *PINK1* mutation carriers have a ∼10 year later age at onset (AAO of 43.2 years) than carriers of biallelic mutations (AAO 32.6 years), but were on average still younger than idiopathic PD (iPD) patients (AAO 70 years) used for comparison in the meta-analysis [19]. Furthermore, neuroimaging studies have demonstrated PD-like changes in humans with monoallelic *PINK1*/*PRKN* mutations [16, 46]. However, the possible role of monoallelic pathogenic *PINK1* and *PRKN* mutations in PD remains a matter of vivid debate: a recent study found 0.089% of monoallelic *PINK1* mutation carriers in cases compared to 0.071% in controls in a large population screen (n=376,558)[22]. Another study did not find any associations between *PRKN* variants and risk of PD. Pathogenic and likely-pathogenic heterozygous single nucleotide variants and copy-number variations were less frequent in PD patients (1.52%) compared to controls (1.8%) [50]. These studies suggest that *PINK1*/*PRKN* monoallelic mutations may not contribute to risk, albeit with limited power and no possibility to assess (subtle) signs of parkinsonism in the seemingly healthy controls or without information on the much more common (monoallelic) *PRKN* mutations. Still, when deep clinical phenotyping was applied, patients with PD harboring *PINK1* or *PRKN* monoallelic pathogenic variants were at a significantly higher rate than healthy controls [20].

Biologically, *PINK1* and Parkin are important in regulating mitochondrial quality control. Activation of the *PINK1*/Parkin mitophagy pathway has been shown to result in the selective removal of dysfunctional mitochondria and their damaged DNA (mtDNA) molecules [12, 17, 42]. In a recent study, Parkin-deficient mice were crossed with “mutator mice”, which harbor a proof-reading defective mitochondrial polymerase gamma. In the resulting animals, mtDNA disintegration and impaired mitophagy led to the release of mtDNA molecules into the extracellular space, where they act as damage-associated molecular patterns (DAMPs) triggering inflammatory pathways [41]. However, whether accumulation of mtDNA damage has an influence on the disease manifestation of *PINK1* and *PRKN*-associated PD still needs to be elucidated.

Herein, we present an in-depth characterization of the mitochondrial genome in blood cells as a potential disease marker for genetic PD. We use four cohorts, including two large and homogeneous founder populations (South Tyrolean and Tunisian Arab Berber) carrying *PINK1* and *PRKN* mutations, neuronal models and postmortem brain tissue to assess mitochondrial dynamics as a marker of disease manifestation.

## Materials and Methods (max 3000)

### Study individuals and genetic analysis

Deep mitochondrial sequencing and real-time PCR assays using TaqMan technology were performed on a total of 411 individuals (**Table 1a and S1**). Patients were recruited from Germany, Italy, Tunisia and Serbia fulfilling the UK Brain Bank Criteria for PD. Demographic and basic clinical data from *PINK1* and *PRKN* mutation carriers (biallelic and monoallelic, n=254) are shown in **Table 1b**. The AAO of disease for individuals with *PINK1* mutations, *PRKN* mutations, and iPD is reported in **Table 1a, 1b and S1**. MtDNA was extracted from whole blood using standard protocols for extraction of genomic DNA. *PINK1* and *PRKN* genetic screening was performed with Sanger sequencing and MLPA (multiplex ligation probe-dependent amplification) as previously described[43]. All individuals with PD were negatively screened for other pathogenic variants in known PD genes (SNCA, LRRK2, VPS35, and DJ1) with gene panel sequencing using either the Illumina NextSeq500 or HiSeq4000 sequencers and MLPA analysis. *PINK1* and *PRKN* mutations refer to ‘possibly, probably or definitely’ pathogenic variants and variants of uncertain significance (VUS, e.g. *PINK1* p.G411S, p.N451T) were not included based on the Movement Disorder Society Genetic mutation database (MDSGene) [18]. *PINK1* and *PRKN* variants were labelled as “pathogenic mutations” based on the criteria of ‘possibly, probably or definitely’ pathogenic with clear guidelines from the MDSGene consortium[18, 27]. All individuals provided informed consent prior to their participation. Specific approvals were obtained from the local ethics committees at the Research Ethics Boards of the Universities of Lübeck and Luxembourg and the respective local ethics committees from the cohort centers. Clinical assessment of all patients was performed by movement disorders specialists. Population-matched healthy controls were used in the study. The AAO was self-reported by patients and based on the occurrence of the first motor symptoms. Familial relationships were defined based on self-report. An overview of individuals in the study is depicted in **Figure 1**.

**Table 1a.**
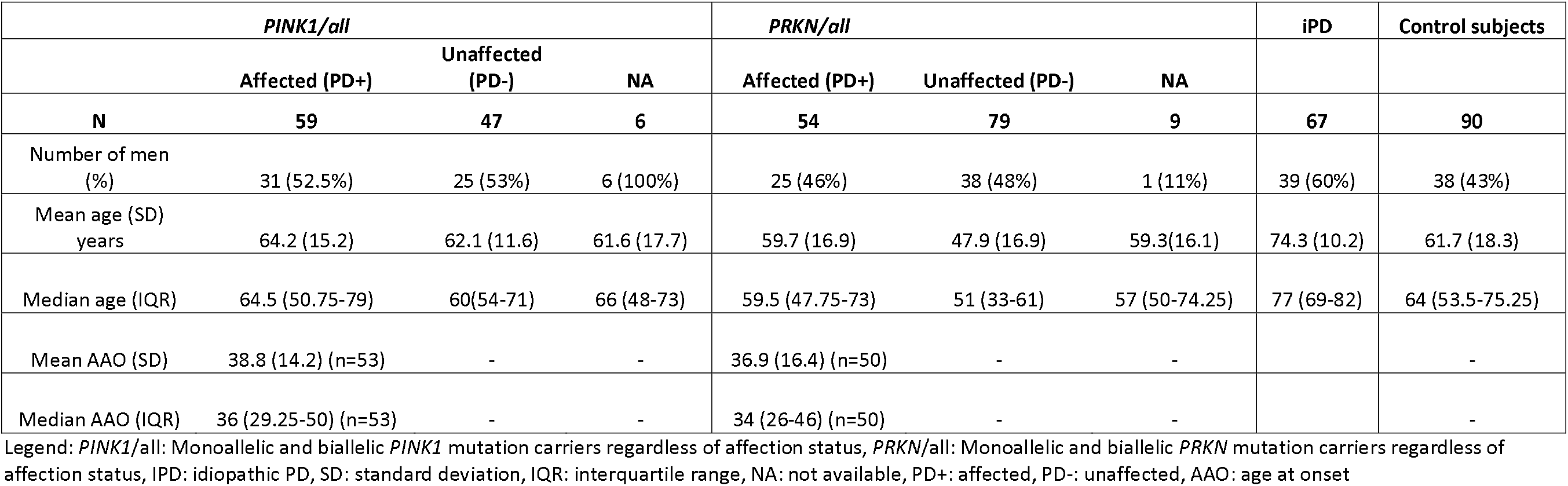
Demographics of all cohorts: individuals with *PINK1/PRKN* mutations, idiopathic Parkinson’s disease and controls.

**Table 1b.** Demographics of cohorts stratified by genotype

|  | *PINK1/all* |  | *PRKN/all* |  |
| --- | --- | --- | --- | --- |
|  | Biallelic (*PINK1*+/+/all) | Monoallelic (*PINK1*+/all) | Biallelic (*PRKN*+/+/all) | Monoallelic (*PRKN*+/all) |
| **N** | 57 | 55 | 27 | 115 |
| Number of men (%) | 29 (51%) | 32 (58%) | 10 (37%) | 54 (47%) |
| Mean age (SD) years | 63.4 (14.6) | 63 (13.1) | 57.3 (17) | 52 (17.9) |
| Median age (IQR) | 63 (51-75) | 61 (54-75) | 58 (48-69) | 54 (36-65.3) |
| Mean AAO (SD) | 37.5 (12.9)<br>(n=51) | 71 (1.4)<br>(n=2) | 35.9 (16.9) (n=28) | 40.2 (15.6)<br>(n=22) |
| Median AAO (IQR) | 36 (29-49.3)<br>(n=51) | 71 (70-71)<br>(n=2) | 32.5 (22.8-46) (n=28) | 39.5 (31.3-50)<br>(n=22) |

Legend: *PINK1*/all: Monoallelic and biallelic *PINK1* mutation carriers regardless of affection status, *PRKN*/all: Monoallelic and biallelic *PRKN* mutation carriers regardless of affection status, *PINK1*+/+: *PINK1* biallelic, *PRKN*+/+: *PRKN* biallelic, *PINK1*+: *PINK1* Monoallelic, *PRKN*+: *PRKN* Monoallelic, SD: standard deviation, IQR: interquartile range, NA: not available, PD+: affected, PD-: unaffected, AAO: age at onset

**Figure 1.**
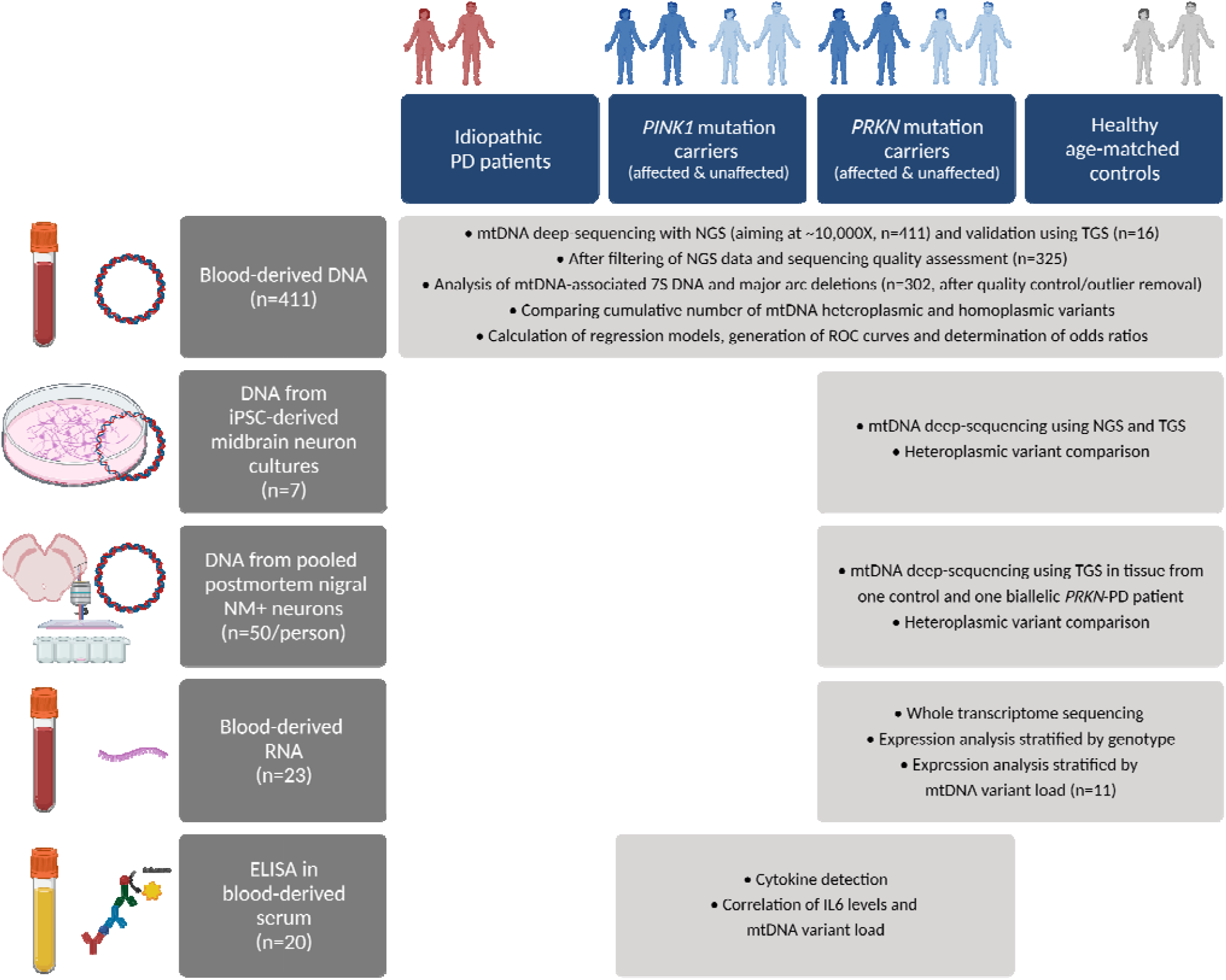
Overview of study. Flow chart showing individuals incorporated in this study, biospecimen, rationale and techniques. mtDNA: mitochondrial DNA, NGS: next-generation sequencing, NM: neuromelanin, TGS: third-generation sequencing.

#### Deep mitochondrial sequencing

##### Illumina NextSeq short-read sequencing

Deep mitochondrial sequencing was performed with bait enrichment with IDT lockdown probes specific for the entire mitochondrial genome to generate libraries of 6-plex or long-range PCR. The two primer sets used for the long-range PCR are described in **Table S2**. Subsequent next-generation sequencing was performed on Illumina, using the NextSeq500 (Illumina, Inc.) to produce 2 × 150bp reads. Raw sequencing reads were converted to standard FASTQ format using bcl2fastq software 2.17.1.14 (Illumina, Inc.). Raw FASTQ files were analyzed with FastQC and aligned with BWA MEM[25] to the mitochondrial reference genome (rCRS) [1]. The resulting SAM files were processed with SAMtools to sorted BAM files [26]. Subsequently, the BAM files were processed with GATK [31] and duplicates removed with MarkDuplicates. Additional quality control was performed with QualiMap 2 [33], and subsequently MultiQC [10] was applied on the resulting reports, including the FastQC reports. The mtDNA-Server [48] successor Mutserve 2 [47] was used for variant calling on the mitochondrial genome to detect homoplasmic and heteroplasmic variants in the sequence data. After additional quality control with Haplocheck [47], we set the minimum heteroplasmy frequency (HF) level to 0.05 [48] and kept all other parameters with the default parameters (per base quality 20, mapping quality 30, alignment score 30), as some samples showed contamination either from different samples or from Nuclear Mitochondrial DNA segments (NUMTs) below that threshold. Mitochondrial genomes with a mean depth of 5,538X (SD:4,919X) coverage were obtained. A cut-off of >1,000X coverage was applied and samples that were contaminated or had an unusually high number of heteroplasmic variants that may be related to NUMTs (>40) were removed (n=86). We defined variants as homoplasmic (>95%) or heteroplasmic (<95% and >5%) based on allele frequency.

#### Oxford Nanopore long-read single-molecule sequencing

To obtain more even coverage across the mtDNA genome and eliminate NUMTs, a long-read sequencing approach was additionally used to detect mtDNA heteroplasmy in blood-derived (n=16) and neuronal-derived DNA (n=10) of *PRKN* mutation carriers and controls. The same two primer sets from Illumina sequencing were used for the long-range PCR. Samples were barcoded and multiplexed, and subsequent sequencing was performed on an R9 flow cell. Mitochondrial genomes with a mean depth of 3433.2X (SD:±2299.9X) coverage were obtained. Base-calling was performed with Guppy v5.0.11 with the super-accurate model. The base-called reads were filtered with Filtlong (v0.2.0) to only include the best 50% of the reads, based on Phred quality scores (q-score), with a minimum read length of 9kb. Then, the reads were trimmed with NanoFilt (v2.5.0). Subsequently, the Nanopore reads were aligned against the mitochondrial genome reference sequence using Minimap2 (v2.17) [24]. The alignments were sorted and indexed with Samtools (v1.3.1). Mutserve2 was used to call variants and the same default parameters were applied in the same fashion as with our short-read sequencing. However, different thresholds for HF levels were set at 0.05, 0.02 and down to 0.01. The per base quality scores allowed for more accurate identification of heteroplasmic variants. All variants from third and second generation sequencing were annotated with a predefined file provided by Mutserve2, with allele frequencies from the 1000 Genomes Project [11], HelixMtDB [4], NUMTs defining variants [7] as well as pathogenicity scores [6, 35].

#### MtDNA deletion and 7S DNA analysis

Real-time PCR assays were performed using TaqMan technology. MtDNA deletion rates were derived from the ratio of the mtDNA-encoded genes NADH:ubiquinone oxidoreductase core subunit 4 and NADH-dehydrogenase 1 (ND4:ND1). ND1 is a gene located in the minor arc of the mitochondrial genome, in an area spared from deletions. ND4 is located in the major arc, in a region commonly affected by somatic deletions. MtDNA transcription initiation was assessed by calculating the 7S DNA:ND1 ratio, as the relative abundance of 7S DNA in the D-Loop region allows for an estimation of the mitochondrial transcription initiation rate[32]. Forward and reverse primers as well as probes coupled with a non-fluorescent quencher described in **Table S2** were used to quantify the concentrations of ND1, ND4, and D-loop, where 7S DNA is integrated during the initial phase of mtDNA transcription. The reactions were performed in 384-well plates in a final reaction volume of 5 µL using a LightCycler480(Roche) as previously described[14]. To minimize inter-experimental variability, reagents for each real-time PCR analysis were pipetted using an automated liquid handler (BioMek FX). A serial dilution of the plasmid p7D1 encoding the target sequence of ND1, ND4 and the D-Loop region was used to quantify the samples. All samples were measured in triplicates. A mixture of all samples was used to normalize across plates. The average and standard deviation of the normalized samples were used for analysis.

#### Generation of iPSC-derived neurons from PRKN-mutant PD patients and controls

iPSCs were generated from fibroblasts of four PD patients (mean age±SD: 57.75±12.03 years; three females and one male) with biallelic *PRKN* mutations (homozygous c.1072Tdel; compound-heterozygous c.1072Tdel+delEx7; compound-heterozygous delEx4+c.924C>T; compound-heterozygous delEx1+c.924C>T) and three age-matched controls (mean age±SD: 45.33±9.74 years, one female and two male) as previously described[44] and cultured in mTeSR1™ complete medium (StemCell Technologies). First, iPSCs were differentiated into small molecule neural precursor cells (smNPCs) using an established protocol[38]. In short, smNPCs were cultured in N2B27 medium: Neurobasal (Gibco)/DMEM-F12 (Gibco) 50:50 and supplemented with 1% B27 without vitamin A (ThermoFisher), 0.5% N2 (Life Technologies), 1% penicillin-streptomycin (ThermoFisher) and 1% 200 mM glutamine (Westburg). During smNPC cultivation, N2/B27 medium was additionally supplemented with 3 µM CHIR 99021 (Sigma), 150 µM ascorbic acid (Sigma) and 0.5 µM purmorphamine (PMA) (Sigma). The medium was changed every other day.

The induction of midbrain neurons began after smNPCs reached the 15^th^ passage. Once ∼75% confluent, cells were detached by subjection to Accutase (Merck Millipore) for 5 min at 37°C and then centrifuged for 3 min at 300 x g at room temperature. Cells were then resuspended and counted via the Countess II FL Automated Cell Counter (ThermoFisher). 750,000 smNPCs per well were seeded onto Matrigel-coated 6-well plates in N2/B27 medium with 1 µM PMA, 200 µM ascorbic acid and 100 ng FGF8 (PeproTech) for 8 days. Next, cells were cultured in N2/B27 medium with 0.5 µM PMA and 200 µM ascorbic acid for two days. For the next 22 days, cells were cultured in N2/B27 medium with 200 µM ascorbic acid, 500 µM dibutyryl-cAMP (Applichem), 1 ng/mL TGF-β3 (Peprotech), 10 ng/mL GDNF (Peprotech) and 20 ng/mL BDNF (PeproTech). The medium was changed every second day. Finally, DNA was prepared from previously characterized iPSC-derived neurons (n=5 million) [44] with the QIAAmp Minikit (Qiagen) and was quantified using standard methods.

#### Laser-microdissected midbrain tissue from PRKN-mutant PD patients and controls

Frozen human postmortem midbrain tissue sections and associated clinical and neuropathological data were supplied by Juntendo University and the Parkinson’s UK brain bank at Imperial College London. One PD patient (age at death: in their 70s, male, compound-heterozygous *PRKN* delEx2 + delEx3) and one control (age at death: in their 70s, male) was used for this analysis. Frozen midbrain blocks were cryosectioned at ∼15 μm thickness in the transverse plane. For the isolation of neuromelanin-positive neurons from the substantia nigra, sections were gradually thawed and fixed with 4% paraformaldehyde for 10 min. After three brief washes in TBST buffer, sections were left to dry on room temperature. Laser capture microdissection (LCM) was performed with the PALM MicroBeam (Zeiss) and isolated neurons were lysed in a buffer (50 mM Tris-HCl, pH 8.5, 1 mM EDTA, 0.5% Tween-20, 200ng/mL proteinase K) for 3hrs at 55°C and 10min at 90°C. In addition, an area without visible neuromelanin deposits surrounding the substantia nigra was cut-out and lysed in the same manner.

#### RNA Ampliseq Transcriptome

RNA was extracted from blood of *PRKN* biallelic and monoallelic mutation carriers (*PRKN*/all) (n=23, mean age±SD: 56.2±13.2, mean AAO±SD: 38.8±18.2) and controls (n=6, mean age±SD: 71.7±9.1) with RNeasy Qiagen Minikit and DNase I digested prior to assessing the concentration, quality and RNA integrity with Agilent 2100 bioanalyzer, RNA LabChip kit and associated software (Agilent). Ampliseq™ whole transcriptome analysis was performed with the Ion Proton (Life Technologies, Inc) with average of 9.96M±1.14M reads. Transcriptome reads were aligned to the GRCh37/hg19 reference genome and analyzed by RNAseq Analysis plugin (V5.0.0.2) using default analysis settings. DESeq2 R package within Bioconductor was used to test for differential expression by use of negative binomial generalized linear models; the estimates of dispersion and logarithmic fold changes incorporate data-driven prior distributions. The non-normalized counts of sequencing reads/fragments were used in DESeq2’s statistical model that accounts for library size differences internally. Differentially expressed genes (DEGs) with nominally-significant p-values (<0.05) and Log2fold (L2F) changes (>|0.2|) were mapped on KEGG pathways with pathview [28].

#### IL6 measurements in serum

To investigate interleukin 6 (IL6) levels in *PINK1*/*PRKN* biallelic and monoallelic mutation carriers (n=20, mean age±SD: 52.9 years ±14.3 years, 9 females), venous blood was collected in serum tubes and processed within one hour. Serum samples were centrifuged at 4°C for 10 min at 3000g, immediately frozen at -80°C and transferred to a certified diagnostic laboratory, where IL6 levels were measured using the Elecsys IL6 assay (Cobax).

#### Statistical Analyses

The affection status (whether the individual was diagnosed with PD), *PINK1*/*PRKN* mutational dosage or heteroplasmic variant load were used as outcome variables in this study. JMP software, Version 10 (SAS Institute Inc, Cary, NC USA) and R, Version 4.1 were used to compare the clinical observations across different groups (i.e. PD vs asymptomatic; AAO; genotypes) and number of heteroplasmic mtDNA variants. For analyses in the blood-derived samples from the cohorts, Kruskal-Wallis tests (followed by post-hoc pairwise comparisons if p<0.05) and Mann-Whitney U-tests were reported. For distinguishing affection status with heteroplasmic mtDNA variants in monoallelic *PINK1*/*PRKN* mutation carriers, multivariable logistic regression models were used to classify predict status by mtDNA variants and age at examination. Then, the corresponding receiving operating characteristic (ROC) curves were reported. We performed one statistical test in the narrow sense to test the specific hypothesis regarding the *PINK1*/*PRKN* affection status with mtDNA heteroplasmic variant load. All other tests were performed exploratorily only, so that reported p-values are not corrected for multiple testing but to be interpreted descriptively or exploratorily. DESeq2 R package within Bioconductor was used to describe differences in gene expression using negative binomial distribution based on estimate variance-mean dependence in count data from high-throughput sequencing assays. The p-values were attained by the Wald test for the gene expression differences. Pearson correlation coefficient was reported for the correlation of IL6 and heteroplasmic variants. For analyses in iPSC-derived neurons, t-tests were performed.

#### Graphical output

All graphics and figures were created with GraphPad Prism and BioRender.com.

## Results

### Deep mitochondrial DNA sequencing with Illumina technology

After stringent filtering for uncontaminated samples, >1,000X coverage, <40 heteroplasmic mtDNA variants, n=325 individuals were included for further analyses. We identified a total 9,799 variants in 325 individuals. The mean number of overall homoplasmic variants was 28.15 (SD: ±15.1) per person and 2.3 (SD: ±2.67) per person for overall heteroplasmic variants.

### Heteroplasmic mtDNA variants are associated with disease manifestation in PINK1/PRKN mutation carriers

We explored whether an association between mtDNA variant burden and PD status exists by comparing the cumulative mtDNA variant load between symptomatic (PD+) and asymptomatic carriers (PD-) of *PINK1* and *PRKN* monoallelic mutation carriers.

This analysis revealed a higher number of heteroplasmic variants in symptomatic (*PINK1*+/*PRKN*+/PD+, n=29), compared to asymptomatic monoallelic carriers (*PINK1*+/*PRKN*+/PD-, n=109) (Mann-Whitney U-test, Z=2.93, p=0.0033, n=138). The resulting area under the curve indicated good discrimination based on low-level mtDNA variants and age at blood drawn for comparison between symptomatic (PD+) and asymptomatic carriers (PD-) of *PINK1* and *PRKN* monoallelic mutation carriers (AUC=0.83, CI:0.74-0.93) **(Figure 2a and b)**.

**Figure 2.**
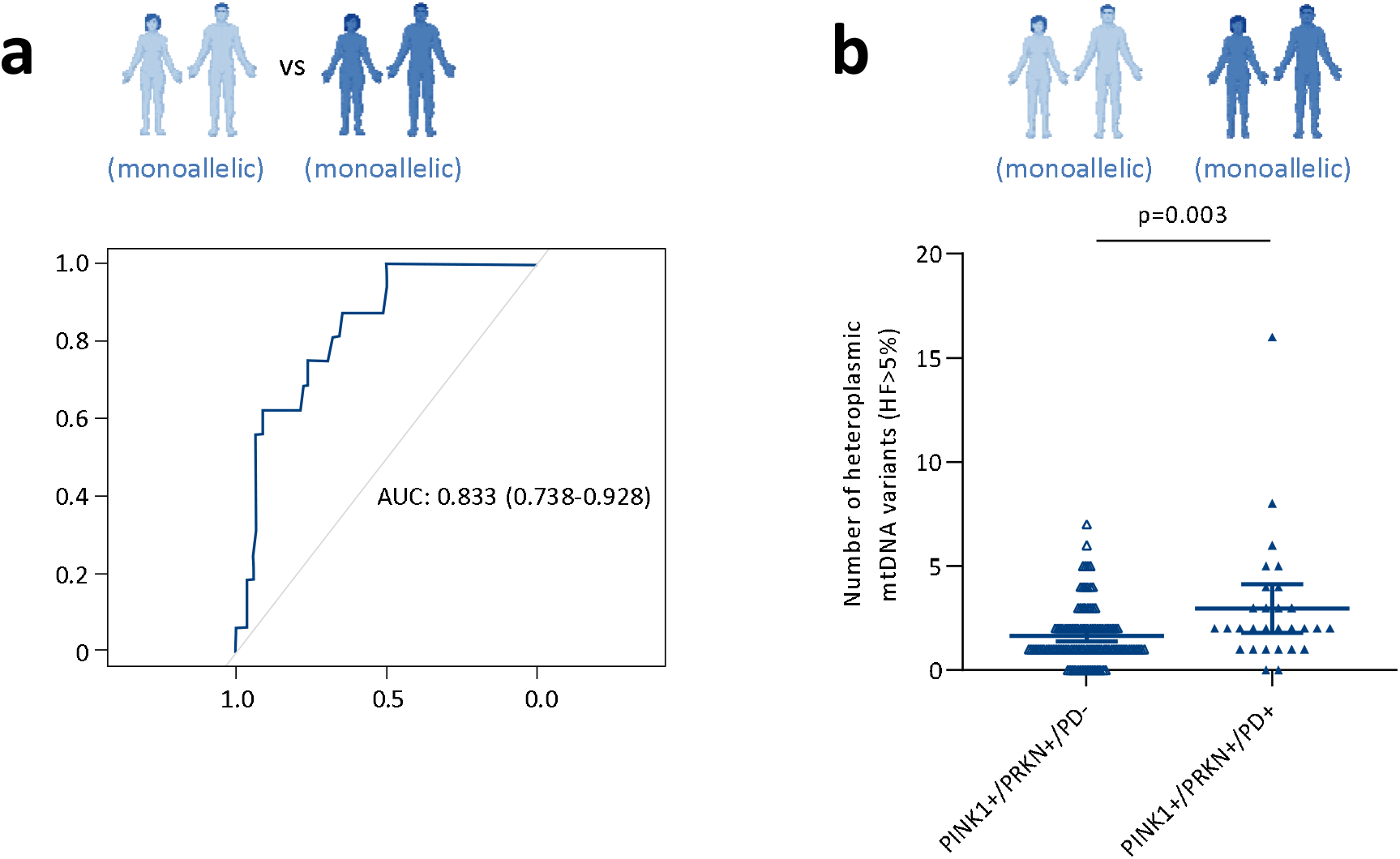
Disease manifestation of *PRKN* and *PINK1* mutations are associated with mtDNA heteroplasmy in blood-derived DNA. **(a)** The sensitivity and specificity of the heteroplasmic mtDNA variants (HF>5%) as a biological marker plus age at examination were assessed with ROC analysis. The area under the curve (AUC) indicated good discrimination for affected (n=29) vs. unaffected (n=109) monoallelic *PRKN/PINK1* mutation carriers (AUC=0.83, CI:0.74-0.93). **(b)** Scatter plot showing the number of heteroplasmic mtDNA variants for affected (n=29) vs. unaffected (n=109) monoallelic *PRKN/PINK1* mutation carriers. Mann-Whitney U-test was performed. Bars indicate means and 95%CI. *PINK1*+:*PINK1* monoallelic mutation; *PRKN*+=*PRKN* monoallelic mutation; PD+: patient with Parkinson’s disease; PD-: individuals without Parkinson’s disease.

By contrast, a comparison of homoplasmic mtDNA variants did not show differences between symptomatic (*PINK1*+/*PRKN*+/PD+, n=29) and asymptomatic *PINK1* or *PRKN* monoallelic mutation carriers (*PINK1*+/*PRKN*+/PD-, n=109) (Mann-Whitney U-test, Z=0.43, p=0.66, data not shown).

### Heteroplasmic mtDNA variants increase with number of mutant PINK1/PRKN alleles

To assess whether the number of *PINK1* or *PRKN* mutations has an impact on the mtDNA variant burden, we compared the heteroplasmic variant load between affected and unaffected individuals with biallelic or monoallelic mutations, idiopathic PD (iPD) patients and controls.

The number of heteroplasmic mtDNA variant load differed between patients with iPD (n=54) and patients with *PINK1*/*PRKN* biallelic or monoallelic mutations (*PINK1*/*PRKN*/PD+, n=78) (Mann-Whitney U-test, p=0.0441) **(Figure 3a)**. Separating patients with monoallelic (*PINK1*+/*PRKN*+/PD+, n=29) and biallelic *PINK1*/*PRKN* mutations (*PINK1*++/*PRKN*++/PD+, n=49), no overall difference was seen to iPD patients (Kruskal-Wallis test, p=0.104) **(Figure 3b)**.

**Figure 3.**
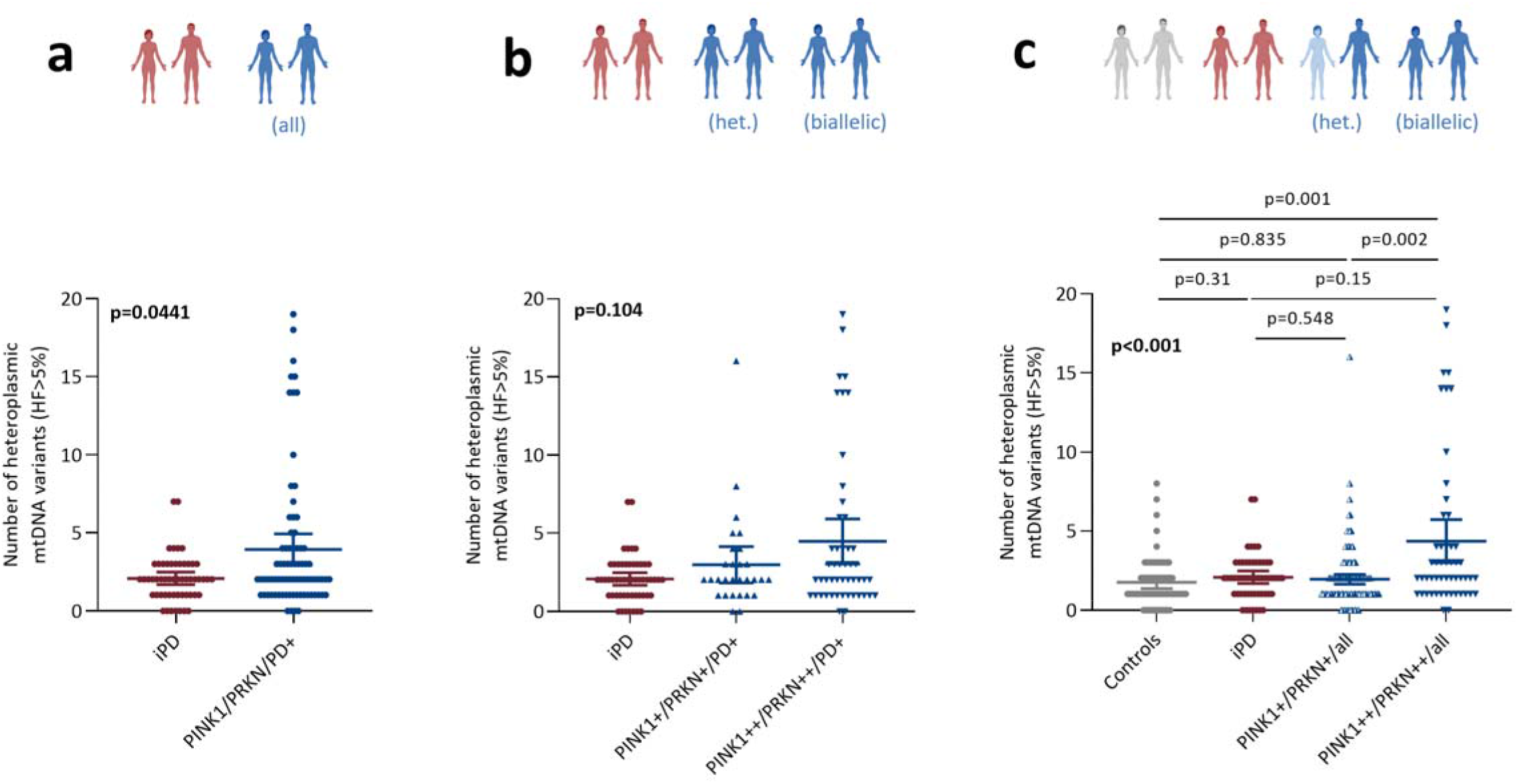
*PRKN*/*PINK1* status is associated with number of heteroplasmic mtDNA variants in blood-derived DNA. Scatter plot showing number heteroplasmic mtDNA variants (HF>5%) for **(a)** idiopathic PD (iPD, n=54) vs *PINK1*/*PRKN*/PD+ (n=78); **(b)** iPD (n=54) vs *PINK1*+/*PRKN*+/PD+ (n=29) vs *PINK1*++/*PRKN*++/PD+ (n=49); **(c)** controls (n=67) vs iPD (n=54) vs all monoallelic mutation carriers (n=150) vs all biallelic mutation carriers (n=52). Mann-Whitney U-test was performed for (a) and Kruskal-Wallis tests for (b) and (c) shown in bold. Post-hoc analyses are not bolded. Bars indicate means and 95%CI. *PINK1*: monoallelic and biallelic *PINK1* mutations; *PRKN*: monoallelic and biallelic *PRKN* mutations; *PINK1*+:*PINK1* monoallelic, *PINK1*++: *PINK1* biallelic mutations; *PRKN*+=*PRKN* monoallelic, *PRKN*++:*PRKN* biallelic mutations; PD+:patient with Parkinson’s disease; all=individuals in study with or without Parkinson’s disease.

Heteroplasmic mtDNA variant load differed between biallelic and monoallelic *PINK1*/ *PRKN* mutation carriers regardless of affection status (*PINK1*/*PRKN*/all) and controls (n=67), higher mtDNA variant load was observed for biallelic compared to monoallelic mutation carriers (Kruskal-Wallis test, p=0.0006) **(Figure 3c)**. Post-hoc pairwise analyses revealed higher heteroplasmic mtDNA variant load for biallelic compared to monoallelic *PINK1*/ *PRKN* mutation carriers (p=0.002) and higher variant load for biallelic *PINK1*/*PRKN*/ mutation carriers compare to controls (p=0.001).

The number of homoplasmic variants also increased with the number of mutant *PINK1*/*PRKN* alleles: mtDNA variants burden differed between patients with iPD (n=54) and *PINK1* or *PRKN* carriers regardless of affection status (n=78) **(Figure S2**, Kruskal-Wallis test, p<0.001). As the homoplasmic mtDNA variants burden should not be influenced by age, we did not perform the age adjustment. These results suggest that heteroplasmic and homoplasmic mtDNA variant load are associated with *PINK1*/*PRKN* genotype.

MtDNA variants were also compared between iPD patients and controls in the same manner. The median values for the number of heteroplasmic variants and homoplasmic variants was not found to differ between iPD patients and controls (Mann-Whitney U-test p=0.0846 and p=0.4668, respectively).

### Quantitative analyses of mtDNA-associated 7S DNA and major arc deletions

To extend our mtDNA variant burden analysis to include large-scale deletions, we investigated ND4:ND1 ratios. After filtering for quality control and removing outliers, n=302 individuals were included for further analyses.

Patients with biallelic *PINK1* or *PRKN* mutations (*PINK1*++/*PRKN*++/PD+; n=57) had more mtDNA deletions than patients with monoallelic *PINK1* or *PRKN* mutations (*PINK1*+/*PRKN*+/PD+; n=25) or iPD cases (n=50) (Kruskal-Wallis test, p=0.009) **(Figure 4a)**. Disregarding the disease status, mtDNA major arc deletions were more abundant in biallelic mutation carriers (n=57) than in monoallelic mutation carriers (n=130) or controls (n=65) (Kruskal-Wallis test, p=0.003) **(Figure 4b)**.

**Figure 4.**
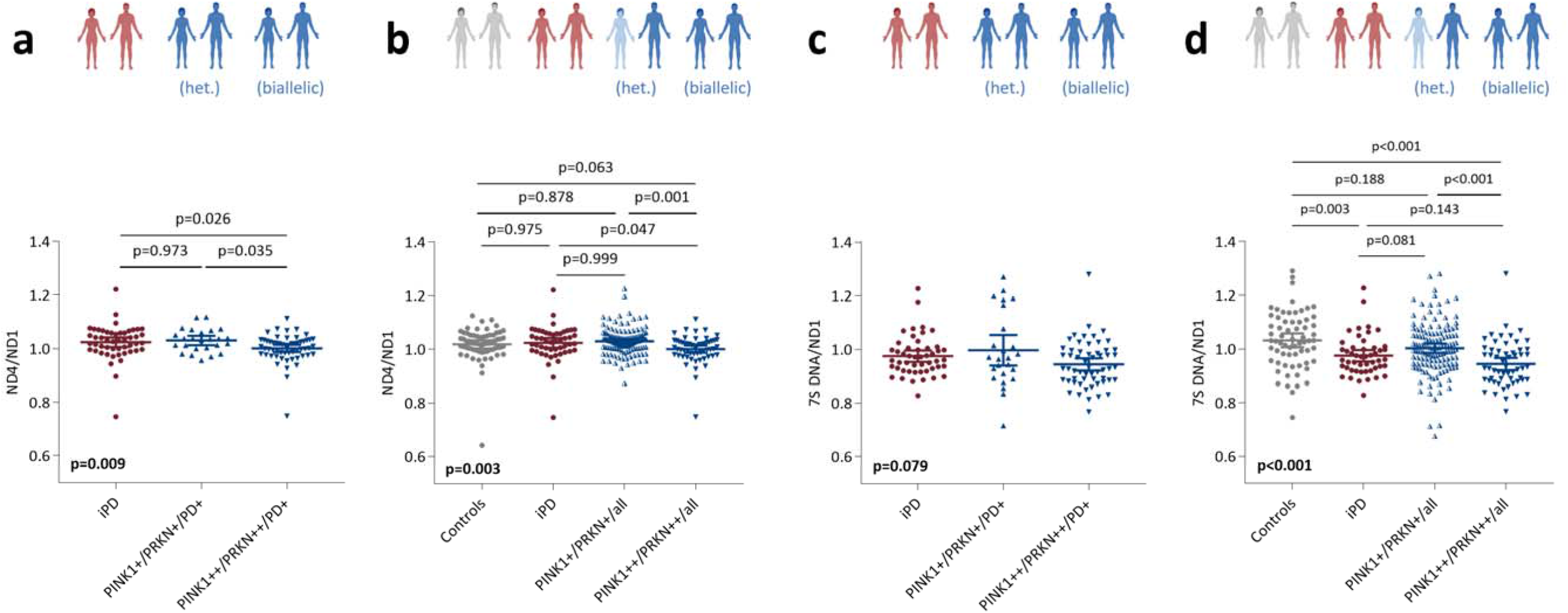
Analysis of mtDNA major arc deletions and mtDNA transcription-associated 7S DNA by real-time PCR in blood-derived DNA. Simultaneous real-time PCR quantification of the mtDNA genes *ND4* relative to *ND1* or detection of 7S DNA relative to *ND1*. Scatter plots showing ND4:ND1 ratios for **(a)** patients with idiopathic PD (iPD, n=50) vs patients with PD due to monoallelic (n=25) or biallelic (n=57) mutations in *PINK1* or *PRKN*. **(b)** Controls (n=65) vs iPD (n=50) vs all *PINK1* or *PRKN* monoallelic mutation carriers (n=130) vs all *PINK1* or *PRKN* biallelic mutation carriers (n=57). Scatter plot showing 7S DNA:ND1 ratios for **(c)** patients with iPD (n=49) vs patients with PD due to monoallelic (n=25) or biallelic (n=57) mutations in *PINK1* or *PRKN*. **(d)** Controls (n=65) vs iPD patients (n=49) vs all monoallelic *PINK1*/*PRKN* mutation carriers (n=131) vs all *PINK1* or *PRKN* biallelic mutation carriers (n=57). Kruskal-Wallis tests were performed and in bold. Post-hoc analyses are not bolded. Bars indicate means and 95%CI; *PINK1*+:*PINK1* monoallelic, *PINK1*++: *PINK1* biallelic mutations; *PRKN*+: *PRKN* monoallelic, *PRKN*++: *PRKN* biallelic mutations; PD+: patient with Parkinson’s disease.

MtDNA maintenance impairments may also extend to changes in transcription and replication of the mitochondrial genome. Events of mtDNA transcription/replication initiation were measured by determining 7S DNA:ND1 ratios **(Figure 4c and 4d)**. Biallelic mutation carriers (*PINK1*++/*PRKN*++/all; n=57) showed lower 7S DNA:ND1 ratios than controls (n=65); a smaller reduction in 7S DNA:ND1 ratios was seen for monoallelic mutation carriers (*PINK1*+/*PRKN*+/all; n=131) compared to controls, and the 7S DNA:ND1 ratios were different between biallelic (*PINK1*++/*PRKN*++/all) and monoallelic (*PINK1*+/*PRKN*+/all) *PINK1* or *PRKN* mutation carriers; lastly, compared to controls, iPD patients (n=49) also showed a similar reduction in 7S DNA:ND1 ratios (Kruskal-Wallis test p<0.001) like biallelic *PINK1* or *PRKN* mutation carriers (*PINK1*++/*PRKN*++/all) **(Figure 4d)**.

### MtDNA variants in iPSC-derived dopaminergic neurons and midbrain tissue with Nanopore sequencing

Next, we tested whether alterations in mtDNA variant load in *PINK1*/*PRKN*-linked PD are a phenomenon restricted to peripheral tissues or whether they can also be observed in patient-derived neurons and midbrain tissue. To assess whether heteroplasmic mtDNA variants are present in neurons, we generated iPSC-derived midbrain neurons from four PD patients with biallelic *PRKN* mutations and three age-matched controls. MtDNA Illumina short-read deep-sequencing of these neuronal cultures revealed an increase in heteroplasmic variants in patient cells (p=0.0389) **(Figure 5a)**. These findings suggests that somatic mtDNA mutation accumulation not only occurs in peripheral tissues but also in iPSC-derived midbrain neurons from PD patients with biallelic *PRKN* mutations.

**Figure 5.**
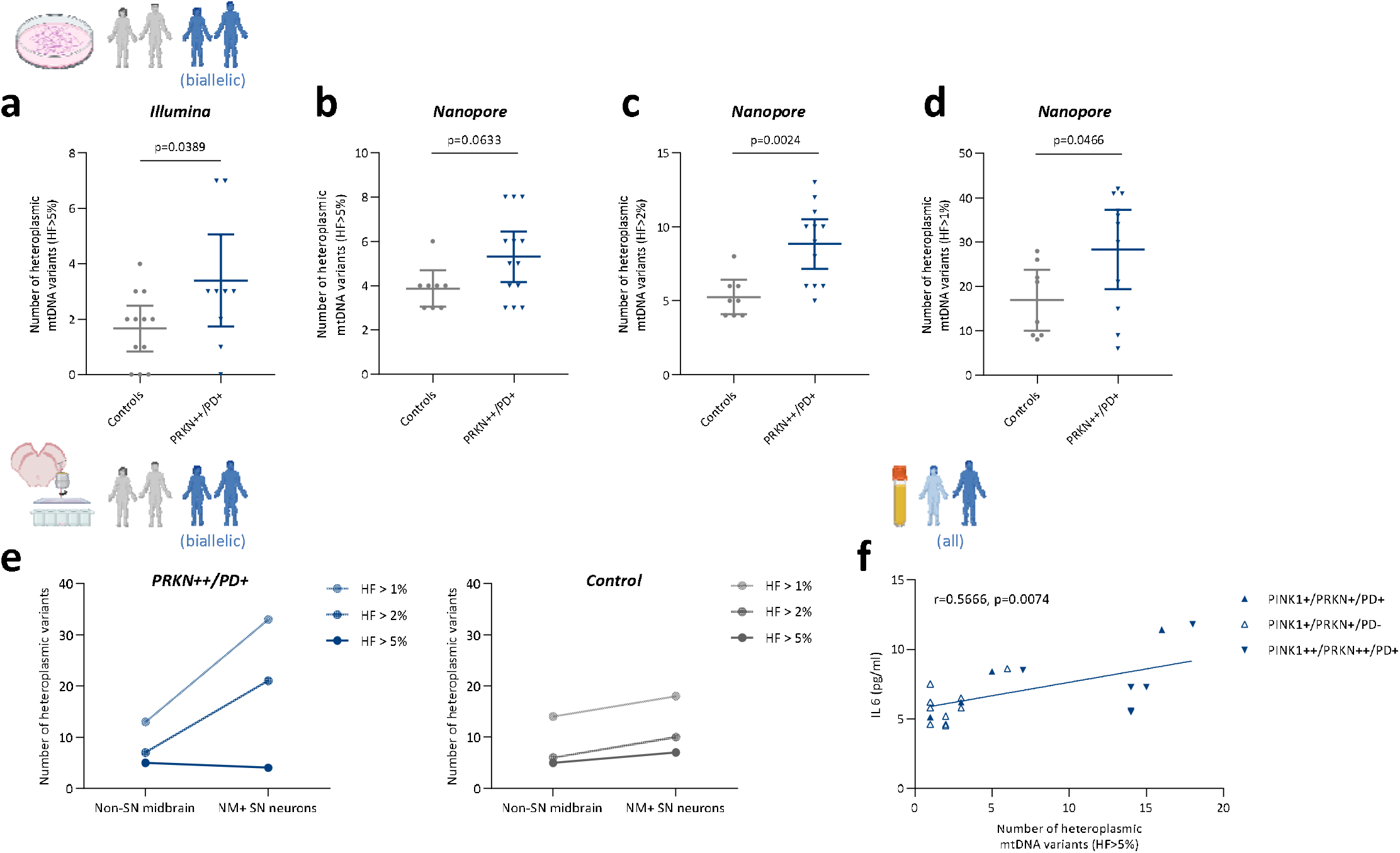
Heteroplasmic mtDNA variants are associated with *PRKN* mutation carriers in iPSC-derived and postmortem neurons and inflammation. Scatter plot showing number of heteroplasmic mtDNA variants in iPSC-derived midbrain neurons from 3-4 differentiations of controls (n=4) and biallelic *PRKN*-PD patients (n=4) performed with **(a)** Illumina short-read sequencing at >5%, **(b)** Nanopore long-read sequencing at >5%, **(c)** Nanopore long-read sequencing at >2%, and **(d)** Nanopore long-read sequencing at >1%. T-tests were performed. *PRKN*++: *PRKN* biallelic mutations; PD+: patients with Parkinson’s disease. **(e)** Line plot showing number heteroplasmic mtDNA variants for control-vs *PRKN*++/PD+-derived nigral laser-microdissected neuromelonin (NM)-positive neurons and non-nigral midbrain tissue at >5%, >2%, and >1%. **(f)** Correlation of serum IL6 and total mtDNA variant load in *PINK1* or *PRKN* monoallelic (n=14) and biallelic (n=6) mutation carriers. Bars indicate means and 95% CI.

Due to limitations in short-read sequencing, we utilized Nanopore long-read sequencing for a more even coverage within amplicons, better mapping quality and single-molecule technology. We first tested the technology in blood-derived *PRKN* monoallelic mutation carriers (n=16 select samples). Nanopore sequencing at >1,000X coverage confirmed 28 out of 32 (87.5%) variants (HF>0.05) in the Illumina short-read dataset. Besides data from our own study, we followed the benchmarking protocol established for Nanopore long-read sequencing in low-frequency variants [29]. When applying 5%, 2% and 1% heteroplasmy thresholds on long-read sequencing data in the neurons, we found that the number of heteroplasmic variants was higher in PD patients with biallelic *PRKN* mutations compared to controls (HF>5%, p=0.0633, HF>2%, p=0.0024, HF>1%, p=0.0466) **(Figure 5b-d)**.

Finally, in light of recently reported limitations of mtDNA studies in iPSC-derived models [45], we compared mtDNA variants in laser-microdissected neuromelanin-positive neurons isolated from postmortem substantia nigra and non-nigral midbrain tissue of a PD patient with biallelic *PRKN* mutations and one control. As we investigated lower HF levels, the difference between the nigral neurons (HF>5%, n=4; HF>2%, n=21; HF>1%, n=33) and the non-nigral midbrain tissue (HF>5%, n=5; HF>2%, n=7; HF>1%, n=13) increases in the *PRKN* mutation carrier. This phenomenon was not as pronounced in nigral neurons (HF>5%, n=7; HF>2%, n=10; HF>1%, n=18) compared to non-nigral midbrain tissue (HF>5%, n=5; HF>2%, n=6; HF>1%, n=14) in the control **(Figure 5e)**.

### MtDNA variant load is associated with differentially-expressed genes (DEGs) in PRKN PD

Recently mtDNA disintegration in *PRKN* KO mutator mice has been linked to mtDNA release and the activation of pro-inflammatory pathways[41]. Thus, we first studied the global expression profile in *PRKN* mutation carriers (*PRKN*/all) and controls from the South Tyrolean cohort with RNA-seq data from available blood samples **(Table S3)**. This analysis revealed 1115 genes with nominally-significant differential expression L2F change >|0.2|(p<0.05). In this set, there was an enrichment for genes in the olfactory transduction pathway (n=22 genes), cytokine-cytokine receptor interaction (n=17 genes) and PI3K-Akt signaling pathway (n=14 genes) **(Table S4)**.

Second, to test whether mtDNA variant load is associated with changes in gene expression, we analyzed the RNA profiles in *PRKN* biallelic and monoallelic patients with more (n=5, mean±SD: 41.1±1.9) or fewer (n=6, mean±SD: 13.3±1.3) homoplasmic mtDNA variants. Still, these individuals were all from the South Tyrolean cohort. This analysis revealed 936 genes with nominally significant differential expression between high and low mtDNA variant load L2F change >|0.2|(p<0.05 before adjustment), 40 genes overlapped with 1115 DEGs comparing *PRKN* mutation carriers to controls. In this set, there was also an enrichment for genes in the olfactory transduction pathway (n=70 genes), the PI3K-Akt signaling pathway (n=17 genes) and the cytokine-cytokine receptor interaction (n=15 genes) **(Table S4b)**. Three genes showed the strongest effects: CD38 (ADP-ribosyl cyclase/cyclic ADP-ribose hydrolase, L2F=-1.13, p=2.64×10^−6^), GSDMA (Gasdermin A, L2F=-1.16, p=4.44×10^−6^), and RNASE1 (Ribonuclease A Family Member 1, L2F=-1.10, p=2.31×10^−5^) **(Figure S3a)**.

### IL6 levels and mtDNA heteroplasmy

Finally, prompted by our finding of altered gene expression in cytokine-cytokine receptor interaction pathways and by the recent literature suggesting a role for proinflammatory signaling in *PINK1*- and *PRKN*-associated PD[30, 41], we next assessed whether the heteroplasmic mtDNA variant load correlated with serum IL6 levels in a subset of *PINK1* and *PRKN* mutation carriers (*PINK1*/*PRKN*/all, n=20), where serum was available. In these individuals, heteroplasmic mtDNA variant load was found to correlate with serum IL6 levels (r=0.57, p=0.0074) **(Figure 5f)**. Similarly, the heteroplasmic mtDNA variant load correlated with serum IL6 in *PINK1* and *PRKN* monoallelic mutation carriers (*PINK1*+/*PRKN*+/all, n=14)(r=0.86, p=8.1×10^−5^) **(Figure S3b)**. Homoplasmic mtDNA variant load did not correlate with serum IL6 levels (p=0.1812).

## Discussion

In this study, we investigated the mtDNA variant burden as a marker of disease state in patients with *PINK1* and *PRKN* mutations. Deep sequencing of blood-derived mtDNA revealed an accumulation of heteroplasmic variants, which are likely to be of somatic origin[34] that could influence the clinical manifestation of *PINK1* or *PRKN* pathogenic mutations. Although the effect size of mtDNA heteroplasmy as a disease marker is relatively low, the ROC curve analyses showed predictive potential after including age (AUC>0.83). Unfortunately, only for a subset of individuals the exact age at blood-draw was available (n=231/325 total individuals). Moreover, the number of affected carriers was too sparse for a continuous AAO trait analysis, and may have been further hampered by the linear association of mtDNA damage and age[21]. However, being mindful of these limitations, the fact that mtDNA heteroplasmy could distinguish between symptomatic and asymptomatic monoallelic carriers of *PINK1* or *PRKN* mutations but not between iPD and controls, still suggests that the effect is specific for individuals with *PINK1* or *PRKN* mutations. As we found that mtDNA heteroplasmy is associated with disease manifestation, we may extrapolate mtDNA heteroplasmic variant load as a penetrance modifier. However, as *PINK1* or *PRKN* monoallelic mutations are still debatable as risk factors of PD, and biallelic mutations are fully penetrant, there are still caveats to this terminology. Interestingly, the analysis of heteroplasmic mtDNA variant load revealed a mutation dosage effect for patients with *PINK1* or *PRKN* mutations. A similar dosage effect was also observed for major arc deletions and 7S DNA. Thus, this finding highlights a mechanistic link between impaired mtDNA maintenance and increased mtDNA variant load. Parkin has been shown to modulate mtDNA transcription by ubiquitination of PARIS, which represses the mitochondrial biogenesis master regulator PGC1-alpha [13]. In addition, our own recent work revealed that a metabolic shift in Parkin-deficient neurons is sensed by SIRT1, which in turn causes a depletion of PGC1-alpha and mtDNA dyshomeostasis [44]. Accordingly, Parkin protects mtDNA from oxidative damage and stimulates mtDNA repair [39].

An increase in mtDNA heteroplasmy likely indicates a development during an individual’s lifetime (somatic variants) that can influence phenoconversion. While somatic mtDNA changes are possibly a direct result of a disturbance in mitochondrial quality control in carriers of *PINK1* or *PRKN* mutations, homoplasmic mtDNA variants are likely to be inherited. The accumulation of homoplasmic variants could be explained by transmission of mtDNA variants across multiple generations over decades in families, a generational conversion of heteroplasmic into homoplasmic variants in populations with *PINK1* or *PRKN* mutations, though this can be influenced by specific mtDNA haplotypes.

To investigate whether the observed mtDNA phenotypes in carriers of *PINK1* or *PRKN* mutations are only a feature detectable in peripheral tissue or whether mtDNA integrity contributes to neurodegeneration in these individuals, we additionally studied neuronal samples. Comparing iPSC-derived midbrain neurons from four PD patients deficient of Parkin and three controls, we confirmed an upregulation of somatic mtDNA mutations in the patient cells. By contrast, recent sequencing analyses in iPSC lines implicated a high mtDNA mutation rate during reprogramming [45]. While this technical limitation alone would not explain the genotype-specific differences in mtDNA mutational load observed here, it motivated us to additionally study postmortem nigral neurons from a biallelic *PRKN*-PD patient and a matched control. Indeed, in laser-microdissected dopaminergic neurons (which were identified based on their neuromelanin deposits) from the *PRKN*-mutant patient, we again detected elevated numbers of somatic mutations. Given that previous deep-sequencing analyses in single postmortem dopaminergic neurons from nigral tissue of iPD patients and controls did not reveal differences in terms of mtDNA somatic mutational load [8], this finding further strengthens the role of Parkin in regulating mtDNA homeostasis. In addition, in line with previous results from iPD patient and control postmortem sections [2, 8], our comparison of the mtDNA status in neuromelanin-positive nigral neurons versus non-nigral midbrain tissue revealed that dopaminergic neurons are particularly vulnerable to this kind of damage. This phenomenon may be explained by the autoxidation properties of dopamine [23], which facilitate the formation of free radicals. ROS, in turn, can cause single- and double-strand breaks, create abasic sites and oxidize purines and pyrimidines in the mitochondrial genome [49].

Apart from its role in mtDNA maintenance, Parkin deficiency has been shown to trigger mtDNA disintegration and dyshomeostasis as a result of impaired mitochondrial clearance [13]. Upon depolarization, oxidative stress or protein misfolding, *PINK1* recruits Parkin to the mitochondria to initiate mitophagy [13, 36]. If mitochondrial clearance fails in the absence of *PINK1* or Parkin, pro-inflammatory signalling is induced. *PRKN* knockout mice with proof reading-deficient mitochondrial polymerase gamma (POLG) as well as PD patients with *PINK1* or *PRKN* mutations showed elevated circulating cell-free mtDNA levels in serum [5], which escapes from cells as consequence of impaired mitophagy. In the extracellular space, mtDNA can act as a damage-associated molecular pattern that is recognized by cGAS/STING and leads to inflammasome activation. Accordingly, *PRKN*-knockout mutator mice and *PINK1* or *PRKN*-mutant PD patients showed an upregulation of the cytokine IL6 in serum [5, 40]. To further investigate the relationship between mtDNA integrity and inflammation, we quantified serum IL6 levels in a small subset of our *PINK1*- and *PRKN* mutation carriers [5, 40]. Remarkably, IL6 concentrations correlated with mtDNA heteroplasmic variant load in these samples. In addition, we observed DEGs enriched in inflammatory pathways when comparing the gene expression profiles of *PRKN* mutation carriers with differential mtDNA variant load. These results concur with the above-mentioned findings in mice and human biosamples[5, 41]. However, cellular studies will be needed to conclusively clarify if inflammation is a downstream effect triggered by accumulation of mtDNA variants and subsequent mtDNA release from damaged mitochondria.

The strength of our study lies in the inclusion of two large founder populations: South Tyroleans and North Africa Arab Berbers, where the frequency of biallelic *PINK1* and *PRKN* variants are higher than in other ethnic groups. These populations provide genetic and environmental homogeneity to increase power for discovery and comprise the majority of our study cohort. Furthermore, our analysis compared specific controls from these founder populations, which also avoids genetic heterogeneity and background bias. However, the disease severity of patients and affection status of currently unaffected carriers may change and longitudinal follow up of our study cohort is warranted. We only had access to a small number of the corresponding mothers of investigated individuals to unequivocally exclude inherited mtDNA variants. In our study, heteroplasmic variants from deep sequencing served as a surrogate marker for somatic mtDNA variation[34]. Using this approach, we provided functional evidence for a role of monoallelic mutations in *PINK1*/*PRKN* as PD risk factors or pathogenic variants of highly reduced penetrance. In addition, we applied single-molecule sequencing to postmortem midbrain tissue and confirmed an upregulation of somatic mutations in nigral dopaminergic neurons from a *PRKN* PD case. Finally, our data provides further evidence for the recently discovered link between the *PINK1*/Parkin mitophagy pathway and inflammation in the context of increased mtDNA heteroplasmy in human samples.

In summary, our findings strengthen the relevance of the *PINK1*/Parkin pathway in maintaining a healthy mitochondrial pool. In addition, our study highlights mtDNA heteroplasmy as a potential disease manifestation marker for *PINK1*/*PRKN* PD. Thus, patient blood-based deep mtDNA sequencing may be considered for genetic counseling and future clinical trials. This is important for monitoring disease onset in asymptomatic carriers, as mtDNA heteroplasmy is reliably measurable. Furthermore, longitudinal prospective studies can aim to prioritize and follow monoallelic mutation carriers for neuroprotective trials before phenoconversion.

## Data Availability

All data produced in the present study are available upon reasonable request to the authors

## Acknowledgments

The authors wish to thank the many patients and their families who volunteered, and the efforts of the many clinical teams involved. Funding has been obtained from the German Research Foundation (“ProtectMove”; FOR 2488, GR 3731/5-1; SE 2608/2-1; KO 2250/7-1), the Luxembourg National Research Fund in the ATTRACT (“Model-IPD”, FNR9631103), NCER-PD (FNR11264123) and INTER programmes (“ProtectMove”, FNR11250962; “MiRisk-PD”, C17/BM/11676395, NB 4328/2-1), the BMBF (MitoPD), the Hermann and Lilly Schilling Foundation, the European Community (SysMedPD), the Canadian Institutes of Health Research (CIHR), Peter and Traudl Engelhorn Foundation. Initial studies in Tunisia on familial parkinsonism were in collaboration with Lefkos Middleton, Rachel Gibson, and the GlaxoSmithKline PD Programme Team (2002-2005). We would like to thank Dr Helen Tuppen from the Welcome Trust Centre for Mitochondrial Research, Newcastle University, UK for providing us with the plasmid p7D1. Moreover, this project was supported by the high throughput/high content screening platform and HPC facility at the Luxembourg Centre for Systems Biomedicine, and the University of Luxembourg.

## Author Contributions

AG, CK and JT designed the study and wrote the manuscript. JT, AG, SD, SI, KW and NO performed the genetic experiments. JT and CVG performed transcriptomic analyses. JT, AH, IK, SD, MW, AG, KL, ELG, and MF performed statistical analyses and data interpretation. JT, HW, PM performed mtDNA Illumina sequencing analysis. JT, TL, SS performed Nanopore sequencing analysis. MB, FH, PB, KKK, SD, AR, VK, AEL, PP, PS, AR, RY, ELG, KL, RY, NB, CK and AG were responsible for clinical recruitment, data acquisition and analyses. AG and CK supervised the study. All authors contributed to critical revision of the manuscript.

## References

1 Andrews RM, Kubacka I, Chinnery PF, Lightowlers RN, Turnbull DM, Howell N (1999) Reanalysis and revision of the Cambridge reference sequence for human mitochondrial DNA. Nat Genet 23: 147 Doi 10.1038/13779

2 Bender A, Krishnan KJ, Morris CM, Taylor GA, Reeve AK, Perry RH, Jaros E, Hersheson JS, Betts J, Klopstock T et al (2006) High levels of mitochondrial DNA deletions in substantia nigra neurons in aging and Parkinson disease. Nat Genet 38: 515–517 Doi 10.1038/ng1769

3 Bloem BR, Okun MS, Klein C (2021) Parkinson’s disease. Lancet 397: 2284–2303 Doi 10.1016/S0140-6736(21)00218-X

4 Bolze A, Mendez F, White S, Tanudjaja F, Isaksson M, Rashkin M, Bowes J, Cirulli ET, Metcalf WJ, Grzymski JJ et al (2019) Selective constraints and pathogenicity of mitochondrial DNA variants inferred from a novel database of 196,554 unrelated individuals. bioRxiv:

5 Borsche M, Konig IR, Delcambre S, Petrucci S, Balck A, Bruggemann N, Zimprich A, Wasner K, Pereira SL, Avenali M et al (2020) Mitochondrial damage-associated inflammation highlights biomarkers in PRKN/PINK1 parkinsonism. Brain 143: 3041–3051 Doi 10.1093/brain/awaa246

6 Castellana S, Biagini T, Petrizzelli F, Parca L, Panzironi N, Caputo V, Vescovi AL, Carella M, Mazza T (2021) MitImpact 3: modeling the residue interaction network of the Respiratory Chain subunits. Nucleic Acids Res 49: D1282–D1288 Doi 10.1093/nar/gkaa1032

7 Dayama G, Emery SB, Kidd JM, Mills RE (2014) The genomic landscape of polymorphic human nuclear mitochondrial insertions. Nucleic Acids Res 42: 12640–12649 Doi 10.1093/nar/gku1038

8 Dolle C, Flones I, Nido GS, Miletic H, Osuagwu N, Kristoffersen S, Lilleng PK, Larsen JP, Tysnes OB, Haugarvoll K et al (2016) Defective mitochondrial DNA homeostasis in the substantia nigra in Parkinson disease. Nat Commun 7: 13548 Doi 10.1038/ncomms13548

9 Dorsey ER, Sherer T, Okun MS, Bloem BR (2018) The Emerging Evidence of the Parkinson Pandemic. J Parkinsons Dis 8: S3–S8 Doi 10.3233/JPD-181474

10 Ewels P, Magnusson M, Lundin S, Kaller M (2016) MultiQC: summarize analysis results for multiple tools and samples in a single report. Bioinformatics 32: 3047–3048 Doi 10.1093/bioinformatics/btw354

11 Genomes Project C, Auton A, Brooks LD, Durbin RM, Garrison EP, Kang HM, Korbel JO, Marchini JL, McCarthy S, McVean GA et al (2015) A global reference for human genetic variation. Nature 526: 68–74 Doi 10.1038/nature15393

12 Gilkerson RW, De Vries RL, Lebot P, Wikstrom JD, Torgyekes E, Shirihai OS, Przedborski S, Schon EA (2012) Mitochondrial autophagy in cells with mtDNA mutations results from synergistic loss of transmembrane potential and mTORC1 inhibition. Hum Mol Genet 21: 978–990 Doi 10.1093/hmg/ddr529

13 Grunewald A, Kumar KR, Sue CM (2019) New insights into the complex role of mitochondria in Parkinson’s disease. Prog Neurobiol 177: 73-93 Doi S0301-0082(18)30065-0

14 Grunewald A, Rygiel KA, Hepplewhite PD, Morris CM, Picard M, Turnbull DM (2016) Mitochondrial DNA Depletion in Respiratory Chain-Deficient Parkinson Disease Neurons. Ann Neurol 79: 366–378 Doi 10.1002/ana.24571

15 Hedrich K, Hagenah J, Djarmati A, Hiller A, Lohnau T, Lasek K, Grunewald A, Hilker R, Steinlechner S, Boston H et al (2006) Clinical spectrum of homozygous and heterozygous PINK1 mutations in a large German family with Parkinson disease: role of a single hit? Arch Neurol 63: 833–838. Doi 10.1001/archneur.63.6.833

16 Hilker R, Klein C, Ghaemi M, Kis B, Strotmann T, Ozelius LJ, Lenz O, Vieregge P, Herholz K, Heiss WD et al (2001) Positron emission tomographic analysis of the nigrostriatal dopaminergic system in familial parkinsonism associated with mutations in the parkin gene. Ann Neurol 49: 367–376

17 Kandul NP, Zhang T, Hay BA, Guo M (2016) Selective removal of deletion-bearing mitochondrial DNA in heteroplasmic Drosophila. Nat Commun 7: 13100 Doi 10.1038/ncomms13100

18 Kasten M, Hartmann C, Hampf J, Schaake S, Westenberger A, Vollstedt EJ, Balck A, Domingo A, Vulinovic F, Dulovic M et al (2018) Genotype-Phenotype Relations for the Parkinson’s Disease Genes Parkin, PINK1, DJ1: MDSGene Systematic Review. Mov Disord 33: 730–741 Doi 10.1002/mds.27352

19 Kasten M, Weichert C, Lohmann K, Klein C (2010) Clinical and demographic characteristics of PINK1 mutation carriers--a meta-analysis. Mov Disord 25: 952–954 Doi 10.1002/mds.23031

20 Klein C, Lohmann-Hedrich K, Rogaeva E, Schlossmacher MG, Lang AE (2007) Deciphering the role of heterozygous mutations in genes associated with parkinsonism. Lancet Neurol 6: 652–662 Doi S1474-4422(07)70174-6

21 Kraytsberg Y, Kudryavtseva E, McKee AC, Geula C, Kowall NW, Khrapko K (2006) Mitochondrial DNA deletions are abundant and cause functional impairment in aged human substantia nigra neurons. Nat Genet 38: 518–520 Doi ng1778

22 Krohn L, Grenn FP, Makarious MB, Kim JJ, Bandres-Ciga S, Roosen DA, Gan-Or Z, Nalls MA, Singleton AB, Blauwendraat C et al (2020) Comprehensive assessment of PINK1 variants in Parkinson’s disease. Neurobiol Aging 91: 168 e161–168 e165 Doi 10.1016/j.neurobiolaging.2020.03.003

23 LaVoie MJ, Hastings TG (1999) Dopamine quinone formation and protein modification associated with the striatal neurotoxicity of methamphetamine: evidence against a role for extracellular dopamine. J Neurosci 19: 1484–1491

24 Li H (2018) Minimap2: pairwise alignment for nucleotide sequences. Bioinformatics 34: 3094–3100 Doi 10.1093/bioinformatics/bty191

25 Li H, Durbin R (2009) Fast and accurate short read alignment with Burrows-Wheeler transform. Bioinformatics 25: 1754–1760 Doi 10.1093/bioinformatics/btp324

26 Li H, Handsaker B, Wysoker A, Fennell T, Ruan J, Homer N, Marth G, Abecasis G, Durbin R, Genome Project Data Processing S (2009) The Sequence Alignment/Map format and SAMtools. Bioinformatics 25: 2078–2079 Doi 10.1093/bioinformatics/btp352

27 Lill CM, Mashychev A, Hartmann C, Lohmann K, Marras C, Lang AE, Klein C, Bertram L (2016) Launching the movement disorders society genetic mutation database (MDSGene). Mov Disord 31: 607–609 Doi 10.1002/mds.26651

28 Love MI, Huber W, Anders S (2014) Moderated estimation of fold change and dispersion for RNA-seq data with DESeq2. Genome Biol 15: 550 Doi s13059-014-0550-8

29 Lüth T, Schaake S, Grünewald A, May P, Trinh J, Weissensteiner H Benchmarking low-frequency variant calling with long-read data on mitochondrial DNA. Frontiers in Genetics: 1096

30 Matheoud D, Cannon T, Voisin A, Penttinen AM, Ramet L, Fahmy AM, Ducrot C, Laplante A, Bourque MJ, Zhu L et al (2019) Intestinal infection triggers Parkinson’s disease-like symptoms in Pink1(-/-) mice. Nature 571: 565–569 Doi 10.1038/s41586-019-1405-y

31 McKenna A, Hanna M, Banks E, Sivachenko A, Cibulskis K, Kernytsky A, Garimella K, Altshuler D, Gabriel S, Daly M et al (2010) The Genome Analysis Toolkit: a MapReduce framework for analyzing next-generation DNA sequencing data. Genome Res 20: 1297–1303 Doi 10.1101/gr.107524.110

32 Nicholls TJ, Zsurka G, Peeva V, Scholer S, Szczesny RJ, Cysewski D, Reyes A, Kornblum C, Sciacco M, Moggio M et al (2014) Linear mtDNA fragments and unusual mtDNA rearrangements associated with pathological deficiency of MGME1 exonuclease. Hum Mol Genet 23: 6147–6162 Doi 10.1093/hmg/ddu336

33 Okonechnikov K, Conesa A, Garcia-Alcalde F (2016) Qualimap 2: advanced multi-sample quality control for high-throughput sequencing data. Bioinformatics 32: 292–294 Doi 10.1093/bioinformatics/btv566

34 Payne BA, Wilson IJ, Yu-Wai-Man P, Coxhead J, Deehan D, Horvath R, Taylor RW, Samuels DC, Santibanez-Koref M, Chinnery PF (2013) Universal heteroplasmy of human mitochondrial DNA. Hum Mol Genet 22: 384–390 Doi 10.1093/hmg/dds435

35 Pereira L, Soares P, Triska P, Rito T, van der Waerden A, Li B, Radivojac P, Samuels DC (2014) Global human frequencies of predicted nuclear pathogenic variants and the role played by protein hydrophobicity in pathogenicity potential. Sci Rep 4: 7155 Doi 10.1038/srep07155

36 Pickrell AM, Huang CH, Kennedy SR, Ordureau A, Sideris DP, Hoekstra JG, Harper JW, Youle RJ (2015) Endogenous Parkin Preserves Dopaminergic Substantia Nigral Neurons following Mitochondrial DNA Mutagenic Stress. Neuron 87: 371–381 Doi 10.1016/j.neuron.2015.06.034

37 Pramstaller PP, Schlossmacher MG, Jacques TS, Scaravilli F, Eskelson C, Pepivani I, Hedrich K, Adel S, Gonzales-McNeal M, Hilker R et al (2005) Lewy body Parkinson’s disease in a large pedigree with 77 Parkin mutation carriers. Ann Neurol 58: 411–422 Doi 10.1002/ana.20587

38 Reinhardt P, Glatza M, Hemmer K, Tsytsyura Y, Thiel CS, Hoing S, Moritz S, Parga JA, Wagner L, Bruder JM et al (2013) Derivation and expansion using only small molecules of human neural progenitors for neurodegenerative disease modeling. PLoS One 8: e59252 Doi 10.1371/journal.pone.0059252

39 Rothfuss O, Fischer H, Hasegawa T, Maisel M, Leitner P, Miesel F, Sharma M, Bornemann A, Berg D, Gasser T et al (2009) Parkin protects mitochondrial genome integrity and supports mitochondrial DNA repair. Hum Mol Genet 18: 3832–3850 Doi 10.1093/hmg/ddp327

40 Sliter DA, Martinez J, Hao L, Chen X, Sun N, Fischer TD, Burman JL, Li Y, Zhang Z, Narendra DP et al (2018) Parkin and PINK1 mitigate STING-induced inflammation. Nature 561: 258–262 Doi 10.1038/s41586-018-0448-9

41 Sliter DA, Martinez J, Hao L, Chen X, Sun N, Fischer TD, Burman JL, Li Y, Zhang Z, Narendra DP et al (2018) Parkin and PINK1 mitigate STING-induced inflammation. Nature: Doi 10.1038/s41586-018-0448-9

42 Suen DF, Narendra DP, Tanaka A, Manfredi G, Youle RJ (2010) Parkin overexpression selects against a deleterious mtDNA mutation in heteroplasmic cybrid cells. Proc Natl Acad Sci U S A 107: 11835–11840 Doi 10.1073/pnas.0914569107

43 Trinh J, Lohmann K, Baumann H, Balck A, Borsche M, Bruggemann N, Dure L, Dean M, Volkmann J, Tunc S et al (2019) Utility and implications of exome sequencing in early-onset Parkinson’s disease. Mov Disord 34: 133–137 Doi 10.1002/mds.27559

44 Wasner K, Smajic S, Ghelfi J, Delcambre S, Prada-Medina CA, Knappe E, Arena G, Mulica P, Agyeah G, Rakovic A et al (2022) Parkin Deficiency Impairs Mitochondrial DNA Dynamics and Propagates Inflammation. Mov Disord: Doi 10.1002/mds.29025

45 Wei W, Gaffney DJ, Chinnery PF (2021) Cell reprogramming shapes the mitochondrial DNA landscape. Nat Commun 12: 5241 Doi 10.1038/s41467-021-25482-x

46 Weissbach A, Konig IR, Huckelheim K, Pramstaller PP, Werner E, Bruggemann N, Tadic V, Lohmann K, Baumer T, Munchau A et al (2017) Influence of L-dopa on subtle motor signs in heterozygous Parkin-and PINK1 mutation carriers. Parkinsonism Relat Disord 42: 95-99 Doi S1353-8020(17)30236-5

47 Weissensteiner H, Forer L, Fendt L, Kheirkhah A, Salas A, Kronenberg F, Schoenherr S (2021) Contamination detection in sequencing studies using the mitochondrial phylogeny. Genome Res: Doi 10.1101/gr.256545.119

48 Weissensteiner H, Forer L, Fuchsberger C, Schopf B, Kloss-Brandstatter A, Specht G, Kronenberg F, Schonherr S (2016) mtDNA-Server: next-generation sequencing data analysis of human mitochondrial DNA in the cloud. Nucleic Acids Res 44: W64–69 Doi 10.1093/nar/gkw247

49 Yakes FM, Van Houten B (1997) Mitochondrial DNA damage is more extensive and persists longer than nuclear DNA damage in human cells following oxidative stress. Proc Natl Acad Sci U S A 94: 514–519 Doi 10.1073/pnas.94.2.514

50 Yu E, Rudakou U, Krohn L, Mufti K, Ruskey JA, Asayesh F, Estiar MA, Spiegelman D, Surface M, Fahn S et al (2021) Analysis of Heterozygous PRKN Variants and Copy-Number Variations in Parkinson’s Disease. Mov Disord 36: 178–187 Doi 10.1002/mds.28299

