## Supplementary data for "Mitochondrial DNA heteroplasmy distinguishes disease manifestation in *PINK1*- and *PRKN*-linked Parkinson’s disease"

**Supplementary Appendix**

This appendix has been provided by the authors to give readers additional information about their work.

**Table S1**. Detailed patient and control subject demographics

|  |  | ***PRKN*** |  |  | ***PINK1*** |  |  | **IPD** | **Controls** |
| --- | --- | --- | --- | --- | --- | --- | --- | --- | --- |
| **Country** |  | **PD+** | **PD-** | **NA** | **PD+** | **PD-** | **NA** |  |  |
| **Germany** | *N* | 20 | 6 | 8 | 7 | 16 | 5 | 46 | 25 |
|  | *Number of men (%)* | 9 (45%) | 3 (50%) | 1 (12.5%) | 2 (28%) | 11 (68.8%) | 5 (100%) | 27 (57%) | 9 (36%) |
|  | *Mean age (SD) years* | 69.1 (12.1) | 65.8 (14.6) | 60.1 (17.2) | 72.7(13.9) | 58 (7.57) | 61.6 (17.7) | 72.7 (10.6) | 67.9 (13.4) |
|  | *Median age (IQR)* | 69 (59-79) | 64.5 (52-79.5) | 61 (49-75) | 79(65.2-80.2) | 59.5 (45.8-59.5) | 66 (48-73) | 75 (64-81) | 66.5 (59.2-81.5) |
|  | *Mean AAO (SD)* | 25 (9.2) | - | - |  | - | - | 58.7 (11.0) | - |
|  | *Median AAO (IQR)* | 26 (16-33) | - | - |  | - | - | 60.5 (51-68) | - |
| **Italy (South Tyrol)** | *N* | 6 | 73 | 1 | - | - | - | - | 25 |
|  | *Number of men (%)* | 4 (67%) | 35(48%) | - | - | - | - | - | 12 (48%) |
|  | *Mean age (SD) years* | 72.5 (13.4) | 46.5 (16.4) | - | - | - | - | - | 40.1 (16.4) |
|  | *Median age (IQR)* | 76 (57.5-83.75) | 48 (30.5-60) | - | - | - | - | - | 42 (23-53) |
|  | *Mean AAO (SD)* | 44.2 (13.2) | - | - | - | - | - | - | - |
|  | *Median AAO (IQR)* | 43 (32.5-56.5) | - | - | - | - | - | - | - |
| **Tunisia** | *N* | 9 | NA | NA | 52 | 31 | - | - | 20 |
|  | *Number of men (%)* | 1 (11%) | - | - | 29 (56%) | 14 (45%) | - | - | 10(50%) |
|  | *Mean age (SD) years* | 61 (13.3) | - | - | 63.2 (15.2) | 64.2 (12.8) | - | - | 69.5 (6.7) |
|  | *Median age (IQR)* | 58 (51-70) | - | - | 62.5 (50.25-75) | 65 (54-76) | - | - | 68.5 (64.2-72.5) |
|  | *Mean AAO (SD)* | 37.1 (15.3) | - | - | 38.9 (14.2) | - | - | - | - |
|  | *Median AAO (IQR)* | 33 (26-45) | - | - | 36 (29.3-50) | - | - | - | - |
| **Serbia** | *N* | 19 | - | - | - | - | - | 21 | 20 |
|  | *Number of men (%)* | 11 (58%) | - | - | - | - | - | 12 (57%) | 7 (35%) |
|  | *Mean age (SD) years* | 49.5 (16.2) | - | - | - | - | - | 80 (6.4) | 72 (10.8) |
|  | *Median age (IQR)* | 47 (35.5-65) | - | - | - | - | - | 81 (75-83) | 73.5 (62-82.3) |
|  | *Mean AAO (SD)* | 42.1 (15.6) | - | - | - | - | - | - | - |
|  | *Median AAO (IQR)* | 43 (27-49.5) | - | - | - | - | - | - | - |
| **Total** | *N* | 54 | 79 | 9 | 59 | 47 | 5 | 67 | 90 |
|  | *Number of men (%)* | 25 (46%) | 38 (48%) | 1 (11%) | 31 (52.5%) | 25 (53%) | 5 (100%) | 39 (60%) | 38 (43%) |
|  | *Mean age (SD) years* | 59.7 (16.9) | 47.9 (16.9) | 59.3 (16.1) | 64.2 (15.2) | 62.1 (11.6) | 61.6 (17.7) | 74.3 (10.2) | 61.7 (18.3) |
|  | *Median age (IQR)* | 59.5 (47.75-73) | 51 (33-61) | 57 (50-74.25) | 64.5 (50.75-79) | 60 (54-71) | 66 (48-73) | 77 (69-82) | 64 (53.5-75.25) |
|  | *Mean AAO (SD)* | 36.9 (16.4) | - | - | 38.8 (14.2) | - | - | - | - |
|  | *Median AAO (IQR)* | 34 (26-46) | - | - | 36 (29.25-50) | - | - | - | - |

Legend: *PINK1*: Monoallelic and biallelic *PINK1* mutation carriers regardless of affection status, *PRKN*: Monoallelic and biallelic *PRKN* mutation carriers regardless of affection status, IPD: idiopathic PD, SD: standard deviation, IQR: interquartile range, NA: not available, PD+: affected, PD-: unaffected, AAO: age at onset.

**Table S2. Primers and probes used in the study**

| ***Mitochondrial sequencing*** |  |
| --- | --- |
| **Name or description** | **Primer sequence** |
| MTL‐F1 (first forward primer) | 5’‐ AAAGCACATACCAAGGCCAC ‐3’ |
| MTL‐F2 (second forward primer) | 5’‐ TATCCGCCATCCCATACATT ‐3’ |
| MTL‐R1 (first reverse primer) | 5’‐ TTGGCTCTCCTTGCAAAGTT ‐3’ |
| MTL‐R2 (second reverse primer) | 5’‐ AATGTTGAGCCGTAGATGCC ‐3’ |
| ***MtDNA deletion and 7S DNA analysis*** |  |
| ND1 forward primer | 5'-CCCTAAAACCCGCCACATCTAC-3' |
| ND1 reverse primer | 5'-GAGCGATGGTGAGAGCTAAGGT-3' |
| ND1 probe | VIC-5'-CCATCACCCTCTACATCACCGCCC-3'-TAMRA |
| ND4 forward primer | 5'-CCATTCTCCTCCTATCCCTCAAC-3' |
| ND4 reverse primer | 5'-CACAATCTGATGTTTTGGTTAAACTATATTT-3' |
| ND4 probe | FAM-5'-CCGACATCATTACCGGGTTTTCCTCTTG-3'-MGB |
| D-loop region forward primer | 5'-CCCACACGTTCCCCTTAAATAA-3' |
| D-loop region reverse primer | 5'-CGTGAGTGGTTAATAGGGTGATAGAC-3' |
| D-loop region probe | ALEXA647-5'-ACATCACGATGGATCAC-3'-MGB |

**Table S3**. Demographics of patients and controls for blood-derived RNA sequencing

| ***PRKN* vs Control** |  |  |  |
| --- | --- | --- | --- |
|  | ***PRKN*** |  | **Control** |
|  | **PD+** | **PD-** |  |
| *N* | 13 | 10 | 6 |
| *Mean age (SD) years* | 59 (SD 17.75) | 52.9 (SD 4.15) | 71.7 (SD 9.97) |
| *Median age (range)* | 53 (43-60) | 63.5 (17-87) | 69.5 (62-84) |
| *Mean AAO (SD)* | 38.8 (SD 18.9) | NA | NA |
| *Median AAO (IQR)* | 37 (11-77) | NA | NA |
| **Heteroplasmic mtDNA load** | **High mtDNA load** |  | **Low mtDNA load** |
| *N* | 5 |  | 6 |
| *Mean age (SD) years* | 56.6 (SD 11.01) |  | 64.2 (SD 13.7) |
| *Median age (range)* | 54 (43-73) |  | 61 (52-87) |
| *Mean AAO (SD)* | NA |  | 15 (NA) |
| *Median AAO (IQR)* | NA |  | 15 (NA) |
| *Number of PRKN monoallelics* | 5 |  | 5 |
| *Number of PRKN biallelics* | 0 |  | 1 |

Legend: *PINK1*: Monoallelic and biallelic *PINK1* mutation carriers regardless of affection status, *PRKN*: Monoallelic and biallelic *PRKN* mutation carriers regardless of affection status, IPD: idiopathic PD, SD: standard deviation, IQR: interquartile range, NA: not available, PD+: affected, PD-: unaffected, AAO: age at onset.

**Table S4a**. Transcriptomic analysis and implemented pathways for all *PRKN* mutation carriers compared to controls

| **Pathways** | **stat.mean** | **set.size** | **p.up** | **p.dn** | **p.val** | **q.val** |
| --- | --- | --- | --- | --- | --- | --- |
| **Olfactory transduction** | -2.00 | 22 | 0.99 | 0.01 | 0.01 | 0.13 |
| **Cytokine-cytokine receptor interaction** | -0.61 | 17 | 0.80 | 0.20 | 0.40 | 0.93 |
| **PI3K-Akt signaling pathway** | -0.63 | 14 | 0.81 | 0.19 | 0.38 | 0.93 |
| Endocytosis | -0.11 | 14 | 0.56 | 0.44 | 0.87 | 0.93 |
| Phagosome | 0.11 | 13 | 0.44 | 0.56 | 0.88 | 0.93 |
| **MAPK signaling pathway** | -0.41 | 12 | 0.71 | 0.29 | 0.57 | 0.93 |
| Ribosome | 1.08 | 11 | 0.08 | 0.92 | 0.16 | 0.93 |
| **Protein processing in endoplasmic reticulum** | -0.46 | 10 | 0.74 | 0.26 | 0.53 | 0.93 |
| **Neuroactive ligand-receptor interaction** | 0.28 | 10 | 0.35 | 0.65 | 0.70 | 0.93 |
| RNA transport | -0.16 | 10 | 0.59 | 0.41 | 0.82 | 0.93 |
| Purine metabolism | -0.09 | 10 | 0.55 | 0.45 | 0.90 | 0.93 |
| Chemokine signaling pathway | -0.07 | 10 | 0.54 | 0.46 | 0.93 | 0.93 |

*bolded pathways are common between Table S4b

**Table S4b**. Transcriptomic analysis and implemented pathways for *PRKN* mutation carriers with higher mutation load and lower mutation load

|  | **stat.mean** | **set.size** | **p.up** | **p.dn** | **p.val** | **q.val** |
| --- | --- | --- | --- | --- | --- | --- |
| **Olfactory transduction** | 1.66 | 70 | 0.01 | 0.99 | 0.02 | 0.25 |
| **Neuroactive ligand-receptor interaction** | 0.55 | 19 | 0.22 | 0.78 | 0.45 | 0.93 |
| **PI3K-Akt signaling pathway** | -0.32 | 17 | 0.67 | 0.33 | 0.66 | 0.93 |
| **Cytokine-cytokine receptor interaction** | 0.061 | 15 | 0.47 | 0.53 | 0.93 | 0.93 |
| Calcium signaling pathway | -0.08 | 12 | 0.54 | 0.46 | 0.91 | 0.93 |
| **Protein processing in endoplasmic reticulum** | -1.14 | 11 | 0.93 | 0.07 | 0.14 | 0.68 |
| **MAPK signaling pathway** | -0.46 | 11 | 0.74 | 0.26 | 0.52 | 0.93 |
| Cell adhesion molecules (CAMs) | 0.33 | 11 | 0.33 | 0.67 | 0.65 | 0.93 |
| Jak-STAT signaling pathway | 0.33 | 10 | 0.33 | 0.67 | 0.65 | 0.93 |
| Toll-like receptor signaling pathway | -0.1 | 10 | 0.56 | 0.44 | 0.89 | 0.93 |

*bolded pathways are common between Table S4a

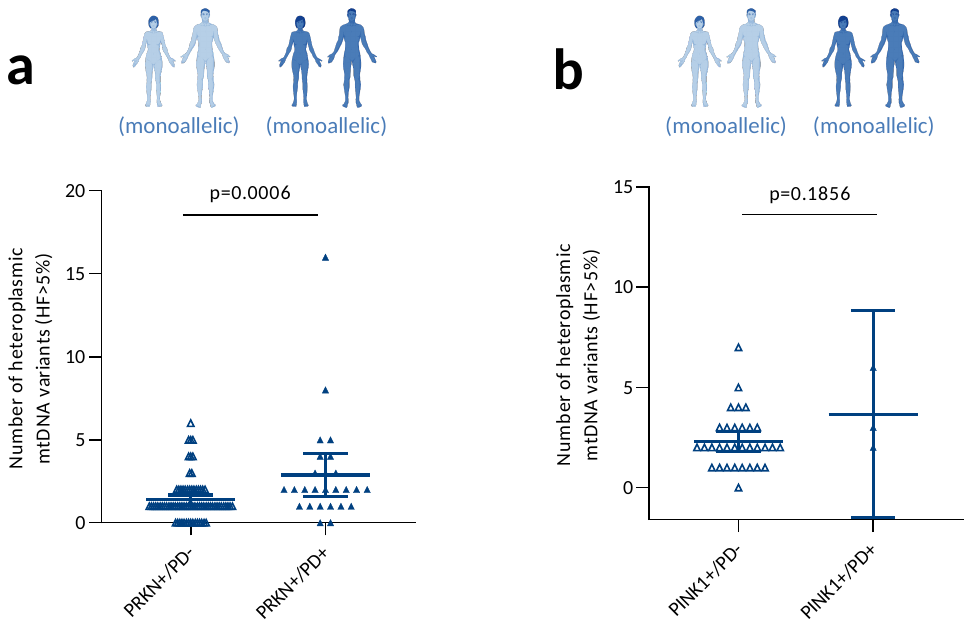

**Figure S1.** **Disease manifestation of *PRKN* and *PINK1* monoallelic mutations are influenced by mtDNA heteroplasmy (HF>5%).** **(a)** Scatter plot showing the number of heteroplasmic variants (HF>5%) in PRKN+/PD- (n=77) vs PRKN+/PD+ (n=26). Bars indicate means and 95%CI. **(b)** Scatter plot showing the number of heteroplasmic variants (HF>5%) in PINK1+/PD- (n=32) vs PINK1+/PD+ (n=3). Bars indicate means and 95%CI. PRKN+=*PRKN* monoallelic; PINK1+=*PINK1* monoallelic; PD+, PD-=participants affected or unaffected with Parkinson’s disease

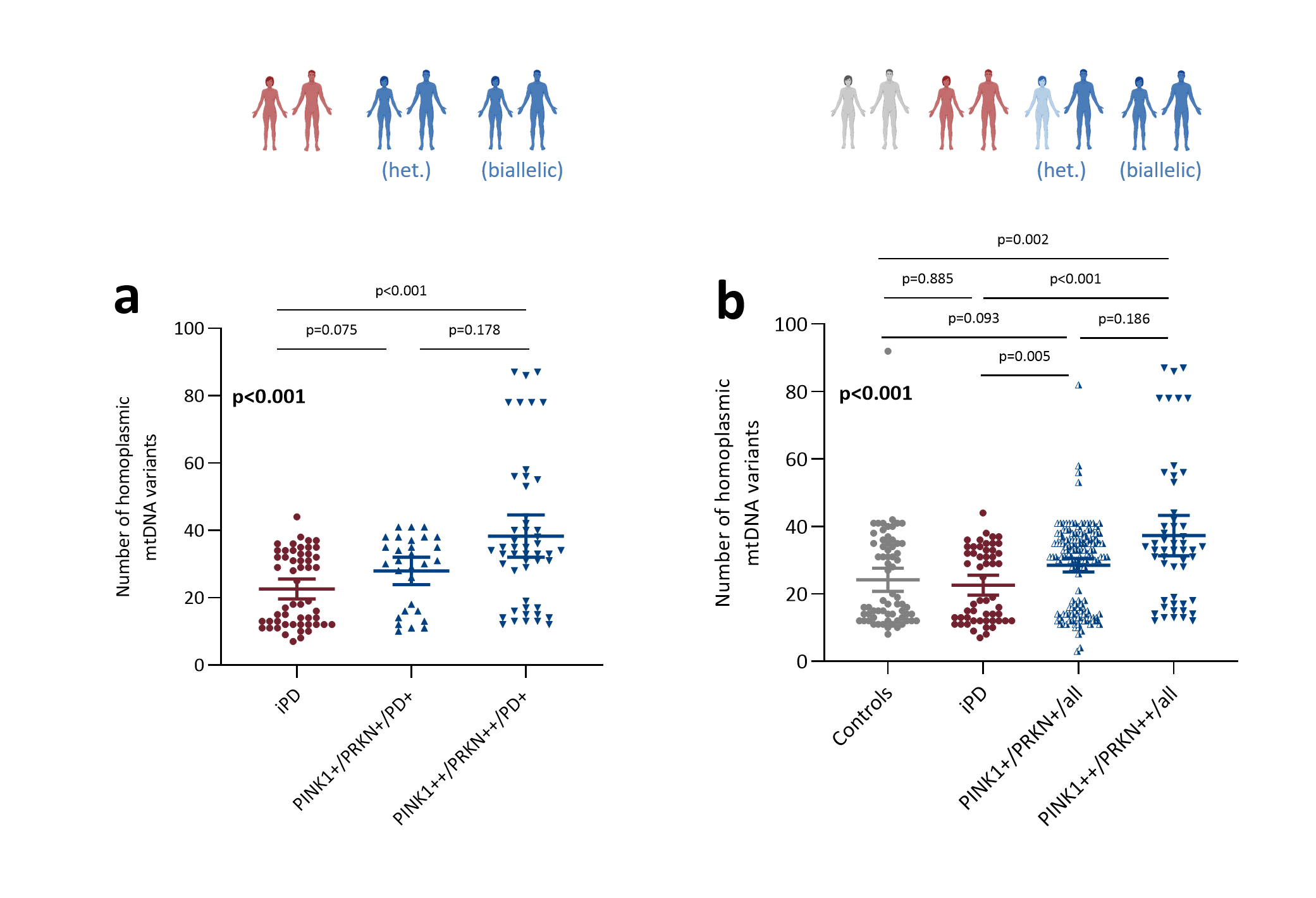

**Figure S2. MtDNA variants and *PINK1/PRKN* genotype status. (a)** Patients only. Scatter plot showing number of homoplasmic mtDNA variants for patients with idiopathic PD (iPD, n=54) vs patients with monoallelic mutations (n=29) vs patients with biallelic *PINK1* or *PRKN* mutations (n=49). **(b)** All individuals in the study. Scatter plot showing number of homoplasmic mtDNA variants for control subjects (n=67) vs iPD (n=54) vs all monoallelic mutation carriers (n=150) vs all biallelic mutation carriers (n=52). Kruskal-Wallis tests were performed and in bold. Post-hoc analyses are not in bold. PINK1+=*PINK1* monoallelic, PINK1++: biallelic mutations detected; PRKN+=*PRKN* monoallelic, PRKN++= *PRKN* biallelic mutations; PD+=patient with Parkinson’s disease, IPD=idiopathic PD.

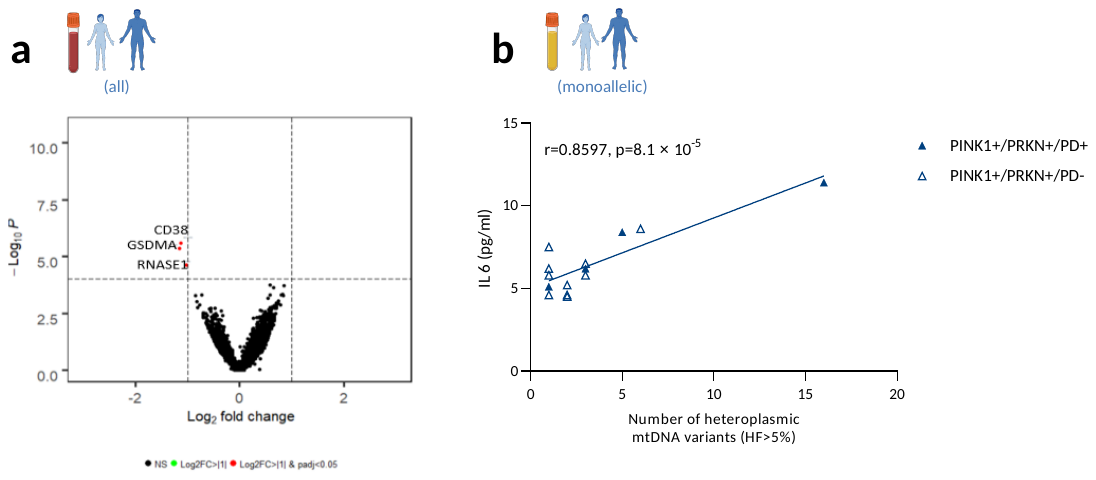

**Figure S3. Differentially expressed genes and IL6 levels in *PRKN* mutation carriers.** **(a)** Volcano plot for transcriptomic analysis of blood comparing *PRKN* mutation carriers with higher and lower mtDNA variant load (n=11), nominating *CD38, GSDMA,* and *RNASE1* as genes of interest. The red dots represents log2fold change > |1|. The dotted lines represent log2fold change > |1| and nominal p-value <10^-5^. **(b)** Correlation of serum IL6 and total mtDNA variant load in affected and unaffected *PINK1* or *PRKN* monoallelic mutation carriers (n=14).

**References**

1.	Trinh J, Lohmann K, Baumann H, Balck A, Borsche M, Bruggemann N, et al. Utility and implications of exome sequencing in early-onset Parkinson's disease. Mov Disord. 2019; **34**(1): 133-7.

2.	Grunewald A, Rygiel KA, Hepplewhite PD, Morris CM, Picard M, Turnbull DM. Mitochondrial DNA Depletion in Respiratory Chain-Deficient Parkinson Disease Neurons. Ann Neurol. 2016; **79**(3): 366-78.

3.	Love MI, Huber W, Anders S. Moderated estimation of fold change and dispersion for RNA-seq data with DESeq2. Genome Biol. 2014; **15**(12): 550.
